# The Influence of Rurality on Self-Reported Physical Therapy Utilization Among Patients with Severe Chronic Back Pain in the United States

**DOI:** 10.1101/2024.12.11.24318576

**Authors:** Kevin H. McLaughlin

**Author notes:** **Corresponding Author:** Kevin McLaughlin, 600 N. Wolfe Street, Baltimore, MD 21287.

## Abstract

**Purpose:** To examine the association of rurality and physical therapy utilization among a nationally representative sample of individuals with severe chronic back pain.

**Methods:** This study utilized a publicly available dataset from the 2019 National Health Information Survey (Adult Sample). Individuals with severe chronic back pain were identified based on survey items examining respondents’ pain frequency and location. Physical therapy utilization was identified based on survey questions asking about pain management strategies utilized over the previous 3 months. Rurality was classified as *large central metro, large fringe metro, medium and small metro*, and *nonmetropolitan/rural*, based on county of residence. Regression was used to examine the association between rurality and physical therapy utilization, while controlling for relevant covariates. National estimates were calculated using provided weighting variables.

**Results:** We identified 2,925 individuals (weighted = 20,468,134) meeting the criteria for severe chronic back pain. We observed that individuals with severe chronic back pain living in nonmetropolitan/rural areas had 34% lower odds of (weighted OR: 0.66, 95% CI: 0.47, 0.92) utilizing physical therapy compared to individuals living in large central metropolitan areas. We did not observe significant differences in the odds of utilizing physical therapy among individuals living in large fringe metropolitan areas or medium-small metropolitan areas.

**Conclusions:** Individuals living in rural parts of the US have lower odds of using physical therapy for management of their severe chronic back pain. Additional research is needed to examine the factors that contribute to these differences and improve access to care.

## Introduction

Back pain is a leading cause of disability globally, affecting more than 500 million individuals annually, with a nearly 20% increase in prevalence over the past 2 decades.^1^ In the US, approximately 80% of adults experience at least 1 episode of back pain during their lifetime, and 25% of adults report back pain that lasted at least 1 day during the past 3 months.^2^ Back pain is also the top non-cancer reason for opioid prescriptions and is the costliest health condition in the US, accounting for an estimated $135 billion in spending, exceeding diabetes, heart disease, and Alzheimer’s disease, and increasing at the second fastest rate of any health condition during the past decade.^3-6^

Physical therapy has been found to be effective for reducing pain and disability among patients with back pain and is recommended as a first line of treatment for back pain in clinical practice guidelines.^7-9^ Physical therapy has also been shown to reduce utilization of opioids and the overall cost of care for patients with back pain.^10-12^ Despite the known benefits of physical therapy, only 7-13% of patients with back pain attend physical therapy. Numerous barriers have been reported that may influence physical therapy utilization for patients with back pain, such as transportation and missed work time.^13-17^ These barriers are likely exacerbated in rural communities where there are 40% fewer physical therapists per capita compared to non-rural areas.^18^ However, physical therapy utilization rates among patients with back pain living in rural communities is unclear. The purpose of this study was to examine the influence of rurality on physical therapy utilization among a nationally representative sample of individuals with severe chronic back pain, using data from the 2019 National Health Information Survey (NHIS).

## Methods

This study utilized publicly available data from the 2019 NHIS (Adult Sample) (https://www.cdc.gov/nchs/nhis/2019nhis.htm) to examine physical therapy utilization among individuals reporting severe chronic back pain. The NHIS is conducted annually by National Center for Health Statistics (NCHS), part of the Centers for Disease Control and Prevention. The NHIS is a cross-sectional household survey, including residents of households and noninstitutional group quarters, such as homeless shelters and group homes ^19^. It does not include individuals without a permanent address, such as active-duty military personnel or those in correctional facilities. The NHIS is conducted face-to-face throughout the year among individuals identified through a geographically clustered sampling technique. The sample is designed in such a way that the overall NHIS sample is nationally representative, allowing researchers to make national estimates based on collected data. Especially important to the completed study, this includes a representative sample of individuals living in rural areas.

### Study Sample

This study included adults participating in the 2019 NHIS who reported severe chronic back pain over the previous 3 months. Severe chronic back pain was operationalized based on participant responses to two survey items. The first item asks participants how often they have experienced pain over the past 3 months: *never, some days, most days*, or *every day*. Individuals responding *most days* or *every day* were be considered to have chronic pain. This approach is used routinely by public health agencies to report the incidence and prevalence of chronic pain in the US^20^ and has been used in several previous studies examining the prevalence of chronic pain using NHIS data.^21,22^ Participants reporting chronic pain are then asked about the severity of pain they experience in multiple body regions, including their back: *not at all, a little, a lot*, or *somewhere in between*.

Participants reporting that their back pain bothers them *a lot* were identified as having severe chronic back pain and were included in the study sample. These same criteria have been used in previous studies to identify patients with severe chronic back pain using 2019 NHIS data. ^22^

### Physical Therapy Utilization

Physical therapy utilization was measured based on participants’ response to a question asking if they utilized physical therapy to manage their pain over the previous 3 months. Participants responding “yes” were classified as having utilized physical therapy.

### Rurality

Rurality was identified based on the 2013 NCHS Urban-Rural Classification Scheme for Counties, which is included in the 2019 NHIS dataset based on participants’ county of residence. Classifications include *large central metro* (1), *large fringe metro* (2), *medium and small metro* (3), and *nonmetropolitan/rural* (4).

### Covariates

We extracted additional participant characteristics from the 2019 NHIS dataset to account for factors other than rurality that might influence participants’ utilization of physical therapy. These included patient age (18-44, 45-64, ≥65 years), insurance coverage (yes/no), education level (less than high school, high school – some college, Associates degree, Bachelor degree – graduate degree) , and income (0-$34,999; $35,000-$49,999; $50,000-$74,999; $75,000-$99,999; ≥$100,000).

### Data Analysis

To account for the complex sampling design of the NHIS that includes stratification and clustering, we utilized survey analysis tools in Stata 17 paired with sampling details (i.e., primary sampling units, strata, weights) included with the NHIS dataset. Summary statistics were used to describe the participants in our sample. Logistic regression was used to examine the association of rurality (independent variable) with receipt of physical therapy (dependent variable). Our statistical model included all previously listed covariates. We considered odds ratios with 95% confidence intervals that did not include 1 to be statistically significant. Weighting variables included in the NHIS dataset were used to create national (weighted) estimates, based on guidance provided by the NCHS.^23^

### Ethical Review

This study was recognized as exempt by the [MASKED] Institutional Review Board.

## Results

We identified 2,925 individuals (weighted = 20,468,134) meeting the criteria for severe chronic back pain. The most commonly represented age group in this study were those between 45-64 years of age (42% of sample, 44% weighted). Individuals included in the study were majority female (60% of sample, 58% weighted), and most commonly identified as White (83% of sample, 82% weighted) and non-Hispanic (91% of sample, 89% weighted) (Table 1).

**Table 1.** Cohort Demographics.

|  | **Sample** | **Weighted** |
| --- | --- | --- |
|  | **N (%)** | **N (%)** |
| Age (years) |  |  |
| 18-44 | 619 (21) | 5,404,616 (26) |
| 45-64 | 1,227 (42) | 9,067,983 (44) |
| ≥65 | 1,079 (37) | 5,995,536 (29) |
| Female | 1,760 (60) | 11,949,283 (58) |
| Race |  |  |
| White | 2,329 (83) | 15,987,900 (82) |
| Black | 328 (12) | 2,276,848 (12) |
| Asian | 36 (1) | 346,824 (2) |
| Native American or Alaskan Native | 34 (1) | 221,381 (1) |
| Other/multi-racial | 60 (2) | 407,732 (2) |
| Unknown | 29 (1) | 244,073 (1) |
| Ethnicity |  |  |
| Hispanic | 262 (9) | 2,176,751 (11) |
| Non-Hispanic | 2,663 (91) | 18,291,384 (89) |

Overall, we observed that 22% (n=703) of our sample and 24% of the weighted sample (n=4,581,674) reported using physical therapy in the previous 3 months. Physical therapy utilization was highest among individuals living in large central metropolitan regions (28% of sample, 27% weighted) and lowest among those living in nonmetropolitan/rural regions (18% of sample, 18% weighted).

We observed that individuals with severe chronic back pain living in nonmetropolitan/rural areas had 34% lower odds (weighted OR: 0.66, 95% CI: 0.47, 0.92) of utilizing physical therapy compared to individuals living in large central metropolitan areas (Table 2). We did not observe significant differences in the odds of utilizing physical therapy among individuals living in large fringe metropolitan areas or medium and small metropolitan areas.

**Table 2.** Physical Therapy Utilization by Rurality.

| **Rurality** | **Utilized Physical Therapy**<br>N (%) |  | **Odds of Utilizing Physical Therapy**<br>OR (95% CI) |  |
| --- | --- | --- | --- | --- |
|  | **Sample** | **Weighted** | **Sample** | **Weighted** |
| Large central metropolitan | 185 (28) | 1,291,158 (27) | *Referent* | *Referent* |
| Large fringe metropolitan | 152 (27) | 975,588 (23) | 0.99 (0.76 – 1.28) | 0.83 (0.61 – 1.14) |
| Medium and small metropolitan | 251 (24) | 1,567,367 (22) | 0.86 (0.69 – 1.1) | 0.79 (0.61 – 1.03) |
| Nonmetropolitan | 115 (18) | 747,562 (18) | 0.67 (0.51 – 0.88) | 0.66 (0.47 – 0.92) |
| Overall | 703 (24) | 4,581,674 (22) | --- | --- |

## Discussion

This is one of the first studies to examine the influence of rurality on physical therapy utilization among individuals with back pain. The results of our study suggest that the odds of using physical therapy as a pain management strategy are 35% lower among individuals with severe chronic back pain living in nonmetropolitan/rural regions compared to those living in large central metropolitan areas. Taken together with previous studies highlighting the differences in physical therapist employment in rural versus nonrural areas,^18,24^ our results suggest there may be rural-urban disparities in access to physical therapy in the US.

Prior to this study, only one other study directly examined the influence of rurality on the utilization of physical therapy among individuals with back pain. ^25^ The previous study utilized a state-wide survey of North Carolina residents to examine healthcare utilization among individuals meeting the National Institutes of Health definition of chronic low back pain, which is similar to the criteria used in the study conducted by this researcher. The investigators of the previous study found no significant differences in physical therapy utilization among individuals with chronic low back pain living in nonmetropolitan versus metropolitan parts of the state. This differs from the results the current study, which identified that individuals living in nonmetropolitan areas were less likely to receive physical therapy for severe chronic back pain. It is possible that these differences are due to the populations surveyed in these studies, as the previous study only included residents of North Carolina, whereas the current study utilized a nationally representative sample of the US adult population. These studies also utilized data collected over 10 years apart (i.e., 2006, 2019).

While the results of this study help to identify differences in physical therapy utilization rates based on rurality, they do not address the underlying causes of these differences. It is possible that observed differences are related to disparities in access, which could be influenced by the fewer number of physical therapists available per capita in rural versus nonrural parts of the country.^18,24^ It is also possible that these differences are related to differences in perceptions of physical therapy among individuals living in rural and nonrural areas. Additional research is needed that examines the factors that influence differences in utilization among individuals living in more or less rural parts of the US in order to inform strategies for addressing differences in utilization.

Strengths of this study include the use of a nationally representative sample of individuals with severe chronic back pain. The primary limitation of this study is its cross-sectional design, which limits our ability to establish causal mechanisms. Additionally, the questions included in the NHIS only ask participants about their use of physical therapy over the previous 3 months. As patients may have received physical therapy for their back pain more than 3 months prior to the time they were surveyed, it is possible that the overall utilization of physical therapy among our study sample is underreported. However, there are no immediate reasons to indicate that this would disproportionately affect responses from individuals living in different regions.

## Conclusion

The odds of utilizing physical therapy are significantly lower among individuals with severe chronic back pain living in rural parts of the US compared to individuals living in large central metropolitan regions. The results of this study suggest there may be rural-urban disparities in access to physical therapy for individuals with severe chronic back pain. Additional research is needed to elucidate the factors that influence these utilization rates and to inform strategies for addressing disparities in access to physical therapy.

## Data Availability

All data used in this study is publicly available.

https://www.cdc.gov/nchs/nhis/documentation/2019-nhis.html

## Acknowledgements

None

## References

1. Vos T, Allen C, Arora M, et al. Global, regional, and national incidence, prevalence, and years lived with disability for 310 diseases and injuries, 1990–2015: a systematic analysis for the Global Burden of Disease Study 2015. The Lancet. October 8, 2016 2016;388(10053):1545–1602. doi:10.1016/S0140-6736(16)31678-6

2. Deyo RA, Mirza SK, Martin BI. Back pain prevalence and visit rates: estimates from U.S. national surveys, 2002. Spine (Phila Pa 1976). Nov 1 2006;31(23):2724–7. doi:10.1097/01.brs.0000244618.06877.cd

3. Dieleman JL, Baral R, Birger M, et al. US Spending on Personal Health Care and Public Health, 1996-2013. JAMA. Dec 27 2016;316(24):2627–2646. doi:10.1001/jama.2016.16885

4. Daubresse M, Chang HY, Yu Y, et al. Ambulatory diagnosis and treatment of nonmalignant pain in the United States, 2000-2010. Med Care. Oct 2013;51(10):870–8. doi:10.1097/MLR.0b013e3182a95d86

5. Smith M, Davis MA, Stano M, Whedon JM. Aging baby boomers and the rising cost of chronic back pain: secular trend analysis of longitudinal Medical Expenditures Panel Survey data for years 2000 to 2007. J Manipulative Physiol Ther. Jan 2013;36(1):2–11. doi:10.1016/j.jmpt.2012.12.001

6. Krebs EE, Gravely A, Nugent S, et al. Effect of Opioid vs Nonopioid Medications on Pain-Related Function in Patients With Chronic Back Pain or Hip or Knee Osteoarthritis Pain: The SPACE Randomized Clinical Trial. JAMA. Mar 6 2018;319(9):872–882. doi:10.1001/jama.2018.0899

7. Chou R, Deyo R, Friedly J, et al. Noninvasive Treatments for Low Back Pain: Comparative Effectiveness Reviews, No. 169. Agency for Healthcare Research and Quality (US). 2016;

8. Chou R, Deyo R, Friedly J, et al. Nonpharmacologic Therapies for Low Back Pain: A Systematic Review for an American College of Physicians Clinical Practice Guideline. Annals of Internal Medicine. 2017-04-04 2017;166(7):493–505. doi:10.7326/M16-2459

9. Fritz JM, Magel JS, McFadden M, et al. Early Physical Therapy vs Usual Care in Patients With Recent-Onset Low Back Pain: A Randomized Clinical Trial. JAMA. 2015 Oct 13 2015;314(14):1459–1467. doi:10.1001/jama.2015.11648

10. Childs JD, Fritz JM, Wu SS, et al. Implications of early and guideline adherent physical therapy for low back pain on utilization and costs. BMC health services research. 2015 Apr 9 2015;15:150. doi:10.1186/s12913-015-0830-3

11. Fritz JM, Childs JD, Wainner RS, Flynn TW. Primary Care Referral of Patients With Low Back Pain to Physical Therapy: Impact on Future Health Care Utilization and Costs. Spine. December 01, 2012 2012;37(25):2114–2121. doi:10.1097/BRS.0b013e31825d32f5

12. Marrache M, Prasad N, Margalit A, et al. Initial presentation for acute low back pain: is early physical therapy associated with healthcare utilization and spending? A retrospective review of a National Database. BMC Health Services Research. 2022;22(1) doi:10.1186/s12913-022-08255-0

13. Carter SK, Rizzo JA. Use of Outpatient Physical Therapy Services by People With Musculoskeletal Conditions. Physical Therapy. May 1, 2007 2007;87(5):497–512. doi:10.2522/ptj.20050218

14. Carvalho E, Bettger JP, Goode AP. Insurance Coverage, Costs, and Barriers to Care for Outpatient Musculoskeletal Therapy and Rehabilitation Services. North Carolina Medical Journal. September 1, 2017 2017;78(5):312. doi:10.18043/ncm.78.5.312

15. Castillo RC, MacKenzie EJ, Webb LX, Bosse MJ, Avery J. Use and perceived need of physical therapy following severe lower-extremity trauma. Archives of physical medicine and rehabilitation. 2005 Sep 2005;86(9):1722–1728. doi:10.1016/j.apmr.2005.03.005

16. Dolot J, Viola D, Shi Q, Hyland M. Impact of Out-of-Pocket Expenditure on Physical Therapy Utilization for Nonspecific Low Back Pain: Secondary Analysis of the Medical Expenditure Panel Survey Data. Physical Therapy. February 1, 2016 2016;96(2):212–221. doi:10.2522/ptj.20150028

17. Dolot J, Hyland M, Shi Q, Kim H-Y, Viola D, Hoekstra C. Factors Impacting Physical Therapy Utilization for Patients With Nonspecific Low Back Pain: Retrospective Analysis of a Clinical Data Set. Physical Therapy. August 31, 2020 2020;100(9):1502–1515. doi:10.1093/ptj/pzaa082

18. Wilson RD, Lewis SA, Murray PK. Trends in the rehabilitation therapist workforce in underserved areas: 1980-2000. The Journal of rural health : official journal of the American Rural Health Association and the National Rural Health Care Association. 2009 Winter 2009;25(1):26–32. doi:10.1111/j.1748-0361.2009.00195.x

19. National Health Interview Survey. Centers for Disease Control and Prevention. 2024. https://www.cdc.gov/nchs/nhis/about_nhis.htm

20. Chronic pain and high-impact chronic pain among U.S. adults, 2019 (National Center for Health Statistics) (2020).

21. Dahlhamer J, Lucas J, Zelaya C, et al. Prevalence of Chronic Pain and High-Impact Chronic Pain Among Adults - United States, 2016. MMWR Morb Mortal Wkly Rep. Sep 14 2018;67(36):1001–1006. doi:10.15585/mmwr.mm6736a2

22. Feldman D, Nahin R. Disability Among Persons With Chronic Severe Back Pain: Results From a Nationally Representative Population-based Sample. The Journal of Pain. 2022/12/01/ 2022;23(12):2144–2154. doi:10.1016/j.jpain.2022.07.016

23. National Center for Health Statistics. National Health Interview Survey - 2019 Survey Description. 2020. https://ftp.cdc.gov/pub/Health_Statistics/NCHS/Dataset_Documentation/NHIS/2019/srvydesc-508.pdf

24. Distribution of U.S. health care providers residing in rural and urban areas. 2013. Accessed 1/18/2024.

25. Goode AP, Freburger JK, Carey TS. The Influence of Rural Versus Urban Residence on Utilization and Receipt of Care for Chronic Low Back Pain. The Journal of Rural Health. 2013;29(2):205–214. doi:10.1111/j.1748-0361.2012.00436.x

